# Non-specific effects of Bacillus Calmette-Guérin - a systematic review and meta-analysis of randomized controlled trials

**DOI:** 10.1101/2022.10.03.22280556

**Authors:** Gerhard Trunk, Maša Davidović, Julia Bohlius

## Abstract

Vaccines induce antigen-specific immunity which provides long-lived protection from the target pathogen. Trials especially from high infectious disease areas indicated that the tuberculosis vaccine Bacillus Calmette-Guérin (BCG) induces in addition non-specific immunity against various pathogens and thereby reduces overall mortality. Although recent trials produced conflicting results, it was suggested that BCG might protect from non-tuberculosis respiratory infections and could be used to bridge time until a specific vaccine against novel respiratory diseases like COVID-19 is available. We performed a systematic search for randomized controlled trials (RCT) published between 2011 and August 5^th^, 2022 providing evidence about non-specific effects after BCG vaccination, assessed their potential for bias, and meta-analyzed relevant clinical outcomes. We excluded RCTs investigating vaccination with an additional vaccine, unless outcomes from a follow-up period before the second vaccination were reported. Our search identified 15 RCTs including 32160 participants. Vaccination with BCG caused an estimated 49% decrease in risk for respiratory infections (HR 0.51, 95% CI 0.33-0.80) with substantial heterogeneity between trials (I^2^=80%; p<0.00001). There was evidence for a protective effect on all-cause mortality of 22% if follow-up was restricted to one year (HR 0.78, 95% CI 0.63-0.96); we did not find evidence for an effect when we considered longer follow-up (HR 0.87, 95% CI 0.75-1.02). Infection-related mortality after BCG-vaccination was reduced by 33% (HR 0.67; 95% CI 0.46-0.99), mortality for sepsis by 38% (HR 0.62, 95% CI 0.41-0.93). There was no evidence for a protective effect of BCG vaccination on infections of any origin (HR 0.84, 95% CI 0.71-1.00), COVID-19 (HR 0.83, 95% CI 0.61-1.14), sepsis (HR 0.82, 95% CI 0.62-1.07) or hospitalization (HR 1.02, 95% CI 0.92-1.14). According to these results, depending on the setting, vaccination with BCG provides time-limited partial protection against non-tuberculosis respiratory infections and may reduce mortality. These findings underline BCG’s potential 1) in pandemic preparedness against novel pathogens especially in developing countries with established BCG-vaccination programs but limited access to specific vaccines; 2) in reducing microbial infections, antimicrobial prescriptions and thus the development of antimicrobial resistance. There is a need for additional RCTs to clarify the circumstances under which BCG’s non-specific protective effects are mediated.

## Introduction

Bacillus Calmette-Guérin (BCG) is an attenuated bacterial vaccine developed against the respiratory infectious disease tuberculosis. According to the current World Health Organization (WHO) policy, it is mainly used in tuberculosis-endemic countries to protect children against the disease [1,2]. Besides its disease-specific protective effects, several studies indicate that BCG might also protect against non-tuberculosis pathogens [3].

Observational studies and trials conducted in newborns in developing countries reported significant reductions in mortality after BCG vaccination, which cannot be solely explained by protection from tuberculosis [4–6]. However, some trials mainly from high-income countries could not confirm these results [7,8]. The mechanisms behind these non-specific effects (NSE) of BCG are incompletely understood but likely involve multifactorial effects on the innate and adaptive immune response. One described mechanism is trained immunity: a state of innate immune memory that can be induced by vaccination or an infection, leading to epigenetic and metabolic reprogramming of innate immune cells such as monocytes and natural killer cells [9–11]. Moreover, emergency granulopoiesis and heterologous T cell reactivity have been described [12,13].

The sudden occurrence of “Coronavirus disease 2019” (COVID-19), a severe and potentially lethal respiratory disease caused by the novel coronavirus SARS-CoV-2, and its spread as a global pandemic triggered intensive scientific efforts to develop a SARS-CoV-2 specific vaccine, which was perceived as the most effective way out of this pandemic. To bridge time until a SARS-CoV-2 specific vaccine is available, it had been suggested to evaluate BCG’s potential in providing non-specific protection, in particular in people with a high risk for COVID-19 [14]. More than 40 randomized controlled trials (RCTs) with approximately 75000 participants worldwide were initiated. However, the fast development of several COVID-19 specific vaccines and the launch of national COVID-19 vaccination programs severely hampered the recruitment of COVID-19 naïve and non-vaccinated participants for BCG-COVID-19 RCTs [15]. Consequently, these trials are behind schedule, and it is questionable whether the effect of a BCG vaccination alone on COVID-19 can be answered by them.

Although COVID-19 specific vaccines are available by now, there is a need for an assessment of BCG’s NSE and its potential as a bridge-vaccination due to 1) its wide availability especially in developing countries which often suffer from timely access to specific vaccines and medical supplies; 2) its potential to activate the immune system against various pathogens, like emerging vaccine resistant SARS-CoV-2 immune escape mutants or novel pathogens in future pandemics. Moreover, BCG-mediated trained immunity may reduce microbial infections in general, thereby lessening drug prescriptions and the development of antimicrobial resistance; it may also reduce the risk for all-cause mortality.

We aimed to evaluate the effects of BCG-vaccination on the risk for non-tuberculosis respiratory infections, COVID-19, non-tuberculosis infections of any origin, sepsis, mortality, and hospitalizations. For this purpose, we systematically identified RCTs published between 2011 and August 5^th^, 2022, which compared the effects of BCG vaccination against no vaccination, assessed their potential for bias, and quantified the estimated effects by meta-analysis.

## Methods

We performed a systematic review and meta-analysis of RCTs reporting NSE of vaccination with BCG. The study protocol is registered in PROSPERO (CRD 42021255017).

### Data sources and search strategy

A systematic literature search was conducted in four electronic databases (Medline via Pubmed and Ovid, Embase, Cochrane CENTRAL, Living Evidence on COVID-19) and three electronic clinical trial registers (Cochrane COVID-19 Study Register, clinical-trials.gov, and WHO International Clinical Trials Registry Platform) from January 1^st^, 2011 to August 5^th^, 2022 (S1 Table). In addition, we screened the reference lists of included literature and relevant reviews for additional references. Literature published in English was considered. The search strategy is described in detail in S2 Table.

### Study selection and eligibility criteria

We included studies that met the following criteria: 1) RCTs; 2) conducted among children, men and/or women; 3) comparing BCG-vaccination with placebo/no vaccination; 4) reporting NSE after BCG vaccination on non-tuberculosis related respiratory infections, non-tuberculosis related infections of any origin, non-tuberculosis related sepsis, non-tuberculosis related mortality (due to all cause, infectious diseases, respiratory infections, sepsis); non-tuberculosis related hospitalizations (due to all cause, infectious diseases, respiratory infections, sepsis). Trials conducted in participants with interfering comorbidities, e.g. patients with bladder cancer for which BCG is used as a therapeutic option, and trials investigating combinations of interventions were excluded (S3 Table). If trial participants received additional vaccines during the follow-up of the study, we included only the period before the second vaccination. We excluded trials in which the timing of a different second vaccination during the follow-up period was unclear. Two reviewers (GT, MD) independently screened titles and/or abstracts of studies retrieved to identify studies that potentially met the inclusion criteria. Full texts of potentially eligible studies were independently assessed for eligibility by two review authors (GT, MD). In case of disagreement between reviewers over the eligibility of particular studies, a third reviewer (JB) was consulted. We used the Rayyan web application for screening [16].

### Data extraction

Study characteristics and outcome data were recorded by one reviewer (GT) using a standardized data collection form. Another reviewer checked the data (MD). If methods or study design were described in several publications, all publications were used to inform data extraction. If publications with additional analyses for a given trial were available, the publication providing most information was considered.

### Risk of bias assessment

Two reviewers (GT, MD) evaluated independently the risk of bias for each included study using the Cochrane Collaboration’s tool for assessing the risk of bias [17]. Any disagreements over the risk of bias in particular studies were resolved by consultation of a third reviewer (JB). Studies were evaluated based on the following criteria: random sequence generation, allocation concealment, blinding of participants/researchers, blinding of outcome assessment, incomplete outcome data, selective reporting, and other bias due to problems not covered by the previous criteria (S4 Table) [18].

### Data analysis

We assessed the following outcomes: non-tuberculosis respiratory infections; COVID-19; infections of any origin; sepsis; mortality due to all-cause, infectious diseases, sepsis; and hospitalization due to all cause, infectious diseases, and respiratory infections. For every outcome, hazard ratios (HR) with 95% confidence interval (CI) were derived from included trials for vaccinated compared to unvaccinated participants by using direct and indirect methods according to Parmar et al. [19]. For meta-analysis a random effects model was assumed. As sensitivity analysis, we used a fixed effect model. We pooled derived HRs by using the generic inverse variance method. We assessed heterogeneity using Chi^2^-tests and quantified heterogeneity using the I^2^ statistic; a p-value < 0.05 was considered significant [20]. For every outcome with at least eight studies, we evaluated potential bias by visual inspection of funnel plot asymmetry [21]; we assessed potential causes of heterogeneity by stratifying the analysis by age, health status, trial region, method of outcome collection, and follow-up time. Contribution of individual trials to the overall result was analyzed by excluding one study at a time. Review Manager (RevMan, version 5.3.5, 2018) was used for all the analyses.

## Results

### Identification of studies

The literature search identified in total 2318 records. 1807 records were from databases and 511 records from registers. After removal of 618 duplicates, 1700 potentially relevant references and citations describing NSE after BCG vaccination were identified and screened for retrieval. Of these, we excluded 1667 reports based on title and abstract because they did not meet the inclusion criteria. The remaining 33 articles were selected for full-text analysis and evaluated in more detail. Of these, 14 were excluded for the following reasons: four articles did not report relevant outcomes, four articles described trials with a study design that was not relevant, one article was a background article, and in five trials participants received additional vaccines during the follow-up period. The remaining 19 articles reported about 15 trials which met all the inclusion- and exclusion criteria and were included in the systematic review and meta-analysis (Fig 1)[22].

**Fig 1.**
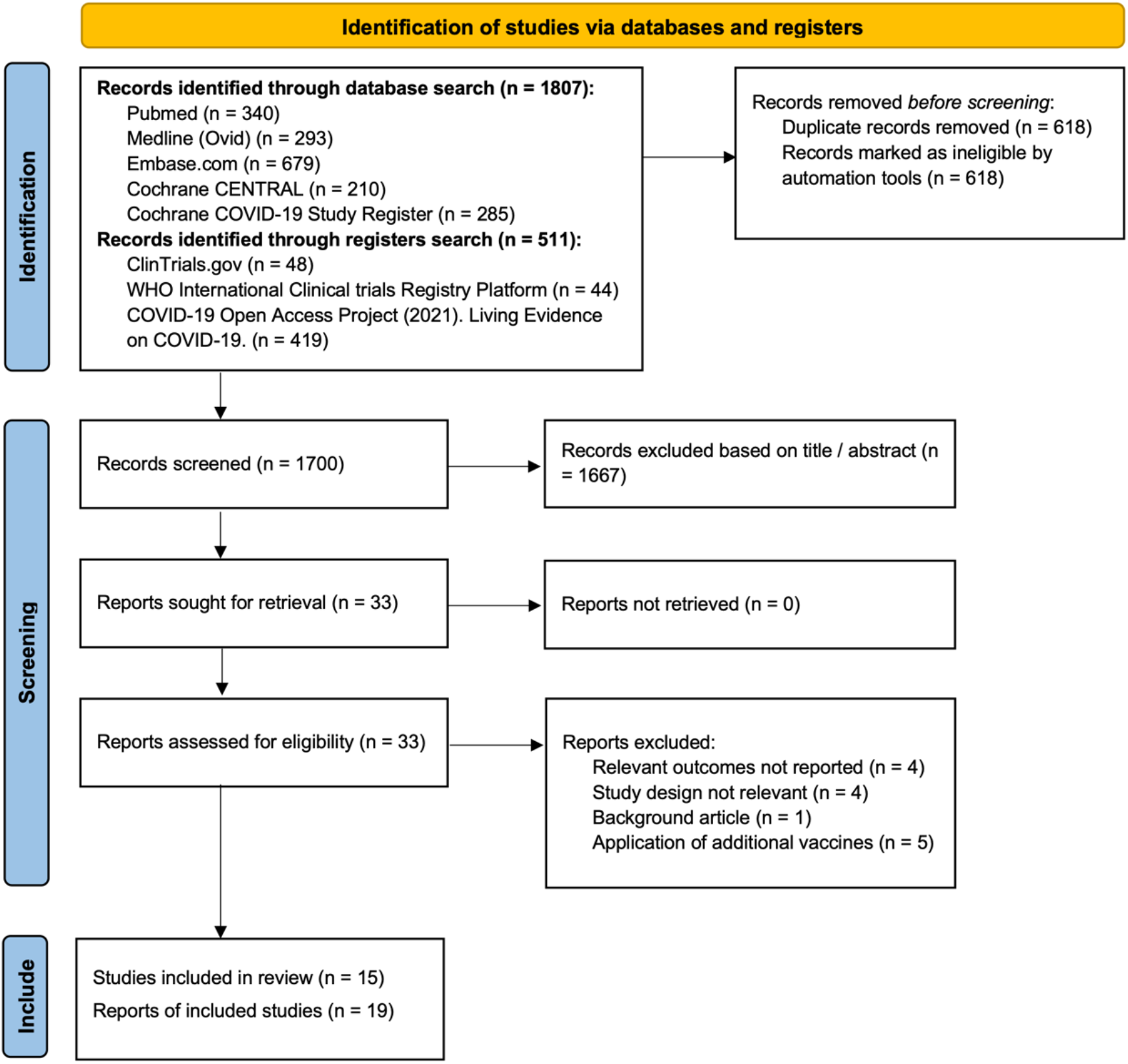
Identification and selection of eligible trials for inclusion in meta-analysis.

### Characteristics of included studies

The 15 identified trials were done in eleven different countries representing low- and high income settings: Guinea-Bissau, Malawi, Uganda, South Africa, Brazil, Indonesia, India, Australia, Denmark, the Netherlands, and Greece (Table 1).

**Table 1.** Characteristics of randomized controlled trials investigating non-specific effects of the BCG vaccine included in meta-analysis. BCG = Bacillus Calmette-Guérin

| Lead author, publication date | Location | Study period | Age group | Intervention (BCG type) | Time of intervention | Control | Follow-up | Total trial participants | Health status |
| --- | --- | --- | --- | --- | --- | --- | --- | --- | --- |
| Aaby et al., 2011<br>Schultz-Buchholzer et al., 2013<br>Schultz-Buchholzer et al., 2018 | Guinea-Bissau | Nov 2002 - Mar 2008 | newborn children | BCG Denmark | immediately after birth | BCG later, i.e. when the child had gained weight or at 6 weeks of age | 12 months | 2320 | low birth weight children < 2500 g |
| Wardhana et al., 2011 | Indonesia | Jun 2009 - Nov 2009 | age 60 - 75 years | BCG weakened living Mycobacterium bovis Pasteur Paris strain mp 1173-P2 produced by PT Biofarma, Bandung, Indonesia | once a month for 3 months in succession | Placebo | 6 months | 34 | healthy individuals |
| Biering-Sorensen et al., 2012<br>Schultz-Buchholzer et al., 2018 | Guinea-Bissau | Nov 2004 - Mar 2008 | newborn children | BCG Denmark | immediately after birth | BCG later, i.e. when the child had gained weight or at 6 weeks of age | 12 months | 104 | low birth weight children < 2500 g |
| Kjærgaard et al., 2016<br>Stensballe et al., 2017<br>Stensballe et al., 2019 | Denmark | Sep 2012 - Jan 2015 | newborn children | BCG Denmark | within 7 days of age | no intervention | 13 months | 4262 | healthy newborns |
| Biering-Sorensen et al., 2017<br>Schultz-Buchholzer et al., 2018 | Guinea-Bissau | Feb 2008 - Sep 2013 | newborn children | BCG Denmark | immediately after birth | BCG later, i.e. when the child had gained weight or at 6 weeks of age | 12 months | 4154 | low birth weight children < 2500 g |
| Nemes et al., 2018 | South Africa | May 2015 - Dec 2016 | age 12 - 17 years | BCG Denmark | revaccination on day 0 (participants were BCG vaccinated in infancy) | Placebo | 24 months | 989 | healthy individuals |
| Jayaraman et al., 2019 | India | Oct 2013 - not reported | newborn children | BCG-Russia | immediately after birth | BCG later, i.e. 28 days after birth | 28 days | 3072 | low birth weight children < 2000 g |
| Giamarellos-Bourboulis et al., 2020 | Greece | Sep 2017 - Nov 2020 | age ≥ 65 years | BCG Denmark | Day of hospital discharge | Placebo | 12 months | 198 | Elderly participants discharged from hospital with various comorbidities |
| Glynn et al., 2021 | Karonga District, northern Malawi | Enrollment of participants: Jan 1986 - Nov 1989 | 3 months - 75 years | BCG (Glaxo-strain) revaccination | revaccination after randomisation | Placebo | Northern area: 1991 - 1994 by active follow-up, Southern area: 2002 - 2018 by demographic surveillance | Northern area: 7389, Southern area: 5616 | Excluded were individuals with past or current leprosy or tuberculosis, severe malnutrition, or other severe illness. |
| Messina et al., 2021 | Australia | Aug 2013 - Sep 2016 | newborn children | BCG Denmark | first 10 days of life | no intervention | 12 months | 1272 | healthy newborns |
| Prentice et al., 2021 | Uganda | Mar 2014 - Jul 2015 | newborn children | BCG Denmark | immediately after birth | BCG later, i.e. 6 weeks after birth | 6 weeks | 560 | healthy infants |
| Tsilika et al., 2022 | Greece | May 2020 - May 2021 | age ≥ 50 years; mean age 69 years | BCG Moscow | day of hospital discharge | Placebo | 6 months | 301 | Elderly participants discharged from hospital with various comorbidities |
| dos Anjos et al., 2022 | Brazil | Aug 2020 - Aug 2021 | adult healthcare workers | BCG Moscow | revaccination after randomisation | no intervention | 180 days | 113 | healthy individuals |
| Doesschate et al., 2022 | Netherlands | Mar 2020 - Apr 2021 | adult healthcare workers | BCG Denmark | (re-)vaccination after randomisation | Placebo | 1 year | 1511 | healthy individuals |
| Upton et al., 2022 | South Africa | Enrollment of participants: May 2020 - Oct 2020 | adult healthcare workers | BCG Denmark | revaccination after randomisation | Placebo | 52 weeks | 265 (per-protocol analysis) | healthy individuals (48.5% with latent tuberculosis) |

The trials included in total 32160 participants. The median number of participants per study was 1131 (interquartile range (IQR) 248 -3343). Seven (47%) trials were conducted in newborn children, one (7%) in adolescent, three in adults (20%) and three trials (20%) in elderly; one (7%) trial included all age groups. Eight (53%) trials [4–6,23–27] mainly included Black participants, two (13%) trials [28,29] mainly Asian participants, and five (33%) trials [7,8,15,30,31] were conducted with participants of mainly Caucasian ethnicity. Nine (60%) trials were conducted in healthy individuals, two (13%) included participants with various comorbidities, and four (27%) were done in low-birth-weight children (<2500g). The primary objectives of 14 (93%) trials [4–8,15,23,25–31] was to evaluate BCG-mediated NSE on mortality, infections of any origin, and respiratory infections; four (27%) trials [15,26,27,31] specifically investigated BCG-mediated NSE on COVID-19; one (7%) trial [24] was conducted to study prevention of M. tuberculosis infection with BCG revaccination. As intervention the BCG Denmark strain was used in ten (67%) trials; one (7%) trial used the BCG Glaxo-strain which is genetically close to the BCG Denmark strain; two trials used the BCG Moscow strain; the remaining two trials used the strains BCG Paris or BCG Russia respectively. Participants were reported to be naïve for BCG in seven trials (47%). Six trials (40%) performed a revaccination with BCG, one of these trials applied BCG once a month for three months in succession [29]. Two (13%) trials did not report previous BCG vaccinations [15,28]. As control intervention placebo was used in seven (47%) trials; three (20%) trials used no intervention as control; five (42%) trials applied in the control group BCG later according to local policies. The follow-up period was in twelve (80%) trials shorter or equal to one year; two (13%) trials had a follow-up between 12 and 24 months; one (7%) trial had follow-up data for four years in one population and for 16 years in another population. Information about application of additional vaccines after BCG within the follow-up period was provided in six (40%) trials [7,8,25–27,31].

### Study quality

Risk of bias assessment of included trials is shown in S4 Table. Two (13%) trials did not report the method of randomization, and six (40%) trials did not describe the method of allocation concealment. Blinding of participants and personnel was unclear in four (27%) trials and was judged to have a high risk of bias in three (20%) trials, in particular due to the collection of participant-reported outcomes without medical diagnosis and visible scar formation after BCG vaccination. In one (7%) trial blinding of assessment was unclear, in another trial (7%) it was not described. Incomplete outcome data due to attrition bias was judged to cause a high risk of bias in five (33%) trials. 14 (93%) trials included an intention-to-treat analysis. One (8%) trial did not inform about the type of analysis.

### Meta-analysis

#### Effects of BCG on non-tuberculosis respiratory infections and COVID-19

We identified eight RCTs including 7392 participants reporting NSE of BCG on non-tuberculosis respiratory infections (Table 2).

**Table 2.**
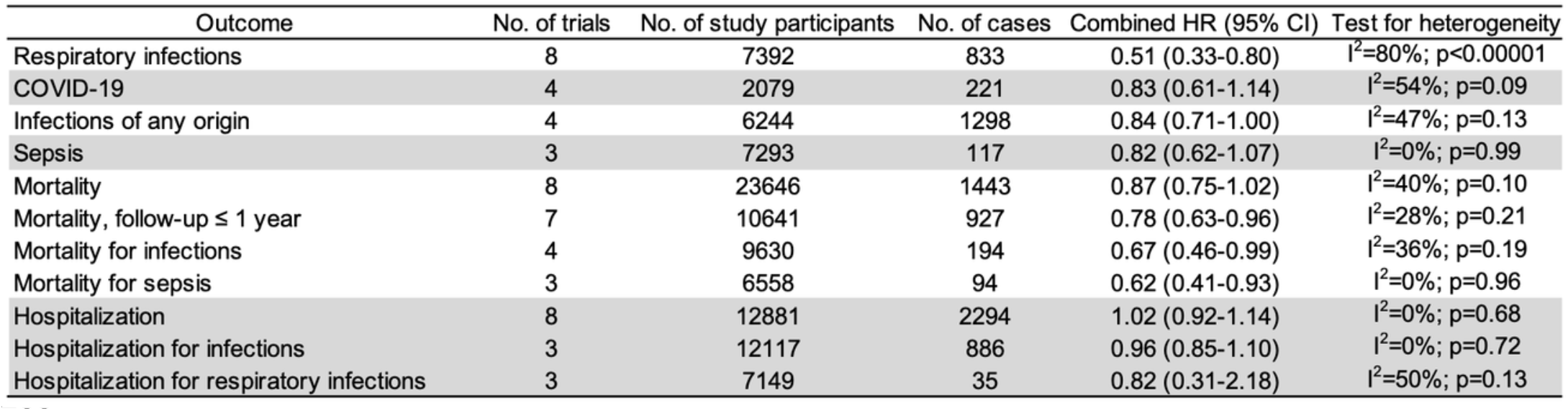
Hazard ratios of respiratory infections, COVID-19, infections of any origin, sepsis, mortality, and hospitalization, according to random-effects meta-analysis of included randomized controlled trials of BCG vaccine that report these outcomes. HR = hazard ratio, CI = confidence interval

Three of them were conducted in healthy newborns (weight 43.8%) [7,8,23], one in adolescents who already received BCG in infancy (weight 18%) [24], one in adults (weight 19.5%) [26], and three in elderly with unknown previous BCG exposure (weight 23.6%) [15,27,28]. Combined HRs from random-effects meta-analyses indicated a beneficial effect of the vaccine on non-tuberculosis respiratory infections (HR 0.51, 95% CI 0.33-0.80); heterogeneity between trials was substantial (I^2^=80%; p<0.00001) (Fig 2A).

**Fig 2.**
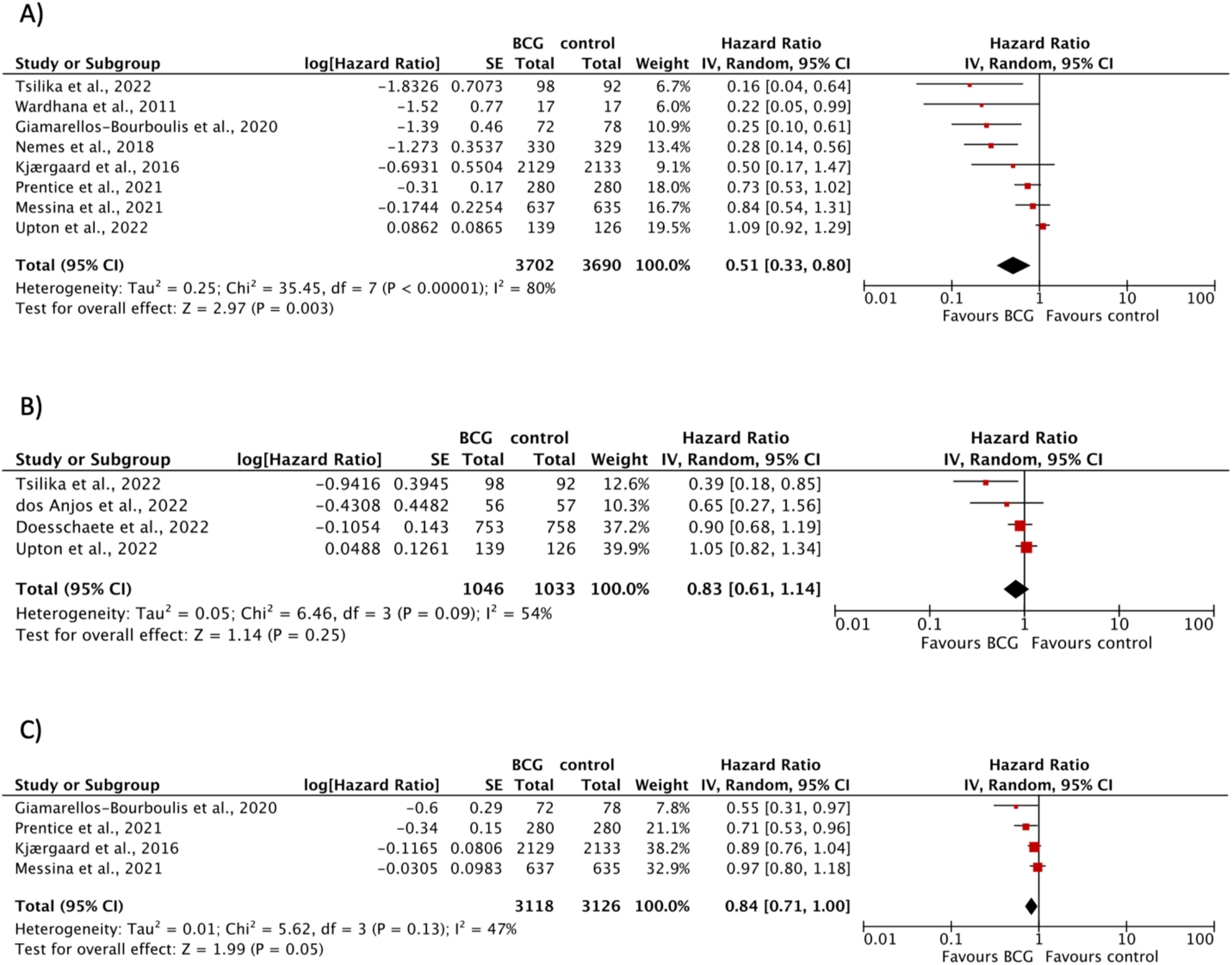
Forest plots of random-effects meta-analysis of BCG trials for A) respiratory infections B) COVID-19 C) infections of any origin. Solid squares represent hazard ratio estimates for the single studies. The size of the squares represents the weight assigned to the individual study in the meta-analysis and is proportional to the inverse variance (IV) of the estimate. Horizontal lines indicate 95% confidence intervals (CI). The diamond shows the 95% CI for the pooled hazard ratios. Values smaller than 1.0 indicate hazard ratios that favor BCG. BCG = Bacillus Calmette-Guérin, SE = standard error

Visual inspection of funnel plots revealed a tendency of small trials to lead to more beneficial intervention effect estimates (S1 Figure). Regarding COVID-19, four RCTs including 2079 participants indicated no evidence for a protective effect after BCG-vaccination (HR 0.83, 95% CI 0.61-1.14) with moderate heterogeneity between trials (I^2^=54%; p=0.09) (Fig 2B). One of them included elderly participants (weight 12.6%) and three were conducted in adult healthcare workers (weight 87.4%). Results remained similar when we used a fixed effect model or excluded one study at a time.

#### Effects of BCG on infections of any origin and sepsis

Four RCTs with a total of 6244 participants investigated the effect of BCG on infections of any origin (Table 2). Three trials were conducted in BCG-naïve healthy newborns (weight 92.2%) and one trial was conducted in elderly (weight 7.8%). There was no evidence for a protective BCG-mediated effect (HR 0.84, 95% CI 0.71-1.00) with moderate heterogeneity between trials (I^2^=47%; p=0.13) (Figure 2C). Exclusion of one study at a time did not extensively change the overall effect. However, applying a fixed effect model resulted in a statistically significant protective effect (HR 0.87, 95% CI 0.78-0.97) (S2 Figure). Regarding sepsis, three RCTs with 7293 participants indicated no evidence for a protective NSE of BCG (HR 0.82, 95% CI 0.62-1.07; I^2^=0%; p=0.99) (Table 2).

#### Effects of BCG on mortality

Eight RCTs with a total of 23646 participants analyzed BCG-mediated effects on all-cause mortality (Table 2). One study followed two populations in two different areas (northern and southern area) during different time-periods with different methods of follow-up and therefore contributed two HRs. There was no evidence for an effect of BCG-vaccination on all-cause mortality (HR 0.87, 95% CI 0.75-1.02) with moderate heterogeneity between the trials (I^2^=40%; p=0.10) (Figure 3A). Restriction to trials with a follow-up of one year excluded one trial [25] which contributed 43.3% weight to the overall analysis and resulted in a statistically significant protective effect (HR 0.78, 95% CI 0.63-0.96) and reduced in-between trial heterogeneity (I^2^=28%; p=0.21) (Fig 3B). Evidence from four trials indicated a protective effect of BCG on infection-related mortality (HR 0.67, 95% CI 0.46-0.99) with moderate heterogeneity between trials (I^2^=36%; p=0.19) (Figure 3C). Mortality for sepsis after BCG vaccination was reported in three trials and resulted in a significantly reduced overall effect estimate (HR 0.62, 95% CI 0.41-0.93) (I^2^=0%; p=0.96) (Figure 3D). Results with fixed effect and random effects models were similar. Only one study reported results about mortality due to respiratory infections [4].

**Fig 3.**
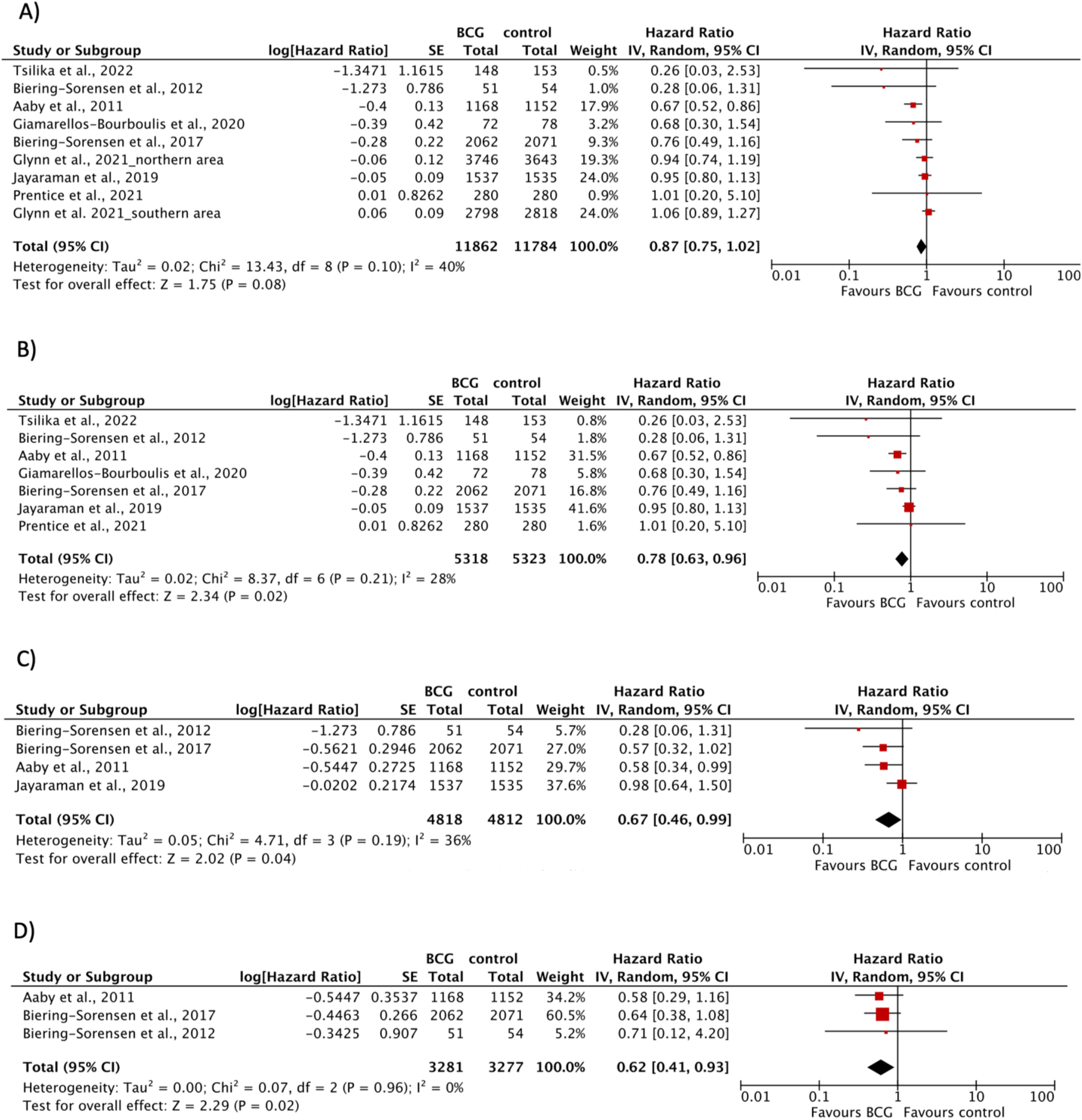
Forest plots of random-effects meta-analysis of BCG trials for A) all-cause mortality B) all-cause mortality with one year follow-up C) mortality for infections D) mortality for sepsis. Solid squares represent hazard ratio estimates for the single studies. The size of the squares represents the weight assigned to the individual study in the meta-analysis and is proportional to the inverse variance (IV) of the estimate. Horizontal lines indicate 95% confidence intervals (CI). The diamond shows the 95% CI for the pooled hazard ratios. Values smaller than 1.0 indicate hazard ratios that favor BCG. BCG = Bacillus Calmette-Guérin, SE = standard error

#### Effects of BCG on hospitalization

We identified eight RCTs including 12881 participants investigating effects of BCG vaccination on all-cause hospitalization (Table 2). The combined HR was 1.02 (0.92-1.14) indicating no evidence for an effect. Trial results were homogenous (I^2^=0%; p=0.68) and the results with fixed effect and random effects models were identical (S3 Figure). Meta-analysis of three trials investigating infectious disease hospitalization and of three trials examining hospitalization for respiratory infections showed no statistically significant improvement after BCG vaccination with combined hazard ratios of 0.96 (95% CI 0.85-1.10) and 0.82 (95% CI 0.31-2.18) respectively (S3 Figure). We found no additional RCTs reporting about hospitalization due to sepsis apart from three RCTs which where meta-analyzed by others before [31].

#### Subgroup analysis

Meta-analysis for the outcomes non-tuberculosis respiratory infections (eight trials), COVID-19 (four trials), infections of any origin (four trials), and mortality (eight trials) showed moderate to substantial heterogeneity between the trials (Table 2). We performed subgroup analyses for outcomes with at least eight trials by stratifying for age, health status, trial region, follow-up period, and method of outcome collection (Table 3). For non-tuberculosis respiratory infections, there was evidence for statistically significant differences between all subgroups (test for subgroup differences: I^2^≥80%; p<0.05), suggesting more pronounced BCG-mediated NSE in non-tuberculosis respiratory infections 1) in adolescents or adults, 2) in low-birth-weight children or morbid participants, 3) in trials conducted in Western Europe or Australia, 4) in RCTs with a follow-up period smaller or equal to six months, 5) and in trials collecting outcome data by medical diagnosis compared to participant-reported data. For mortality, there was evidence for a statistically significant difference between subgroups with different health status, suggesting more pronounced BCG-mediated NSE on mortality in low birth-weight children or morbid participants compared to others.

**Table 3.**
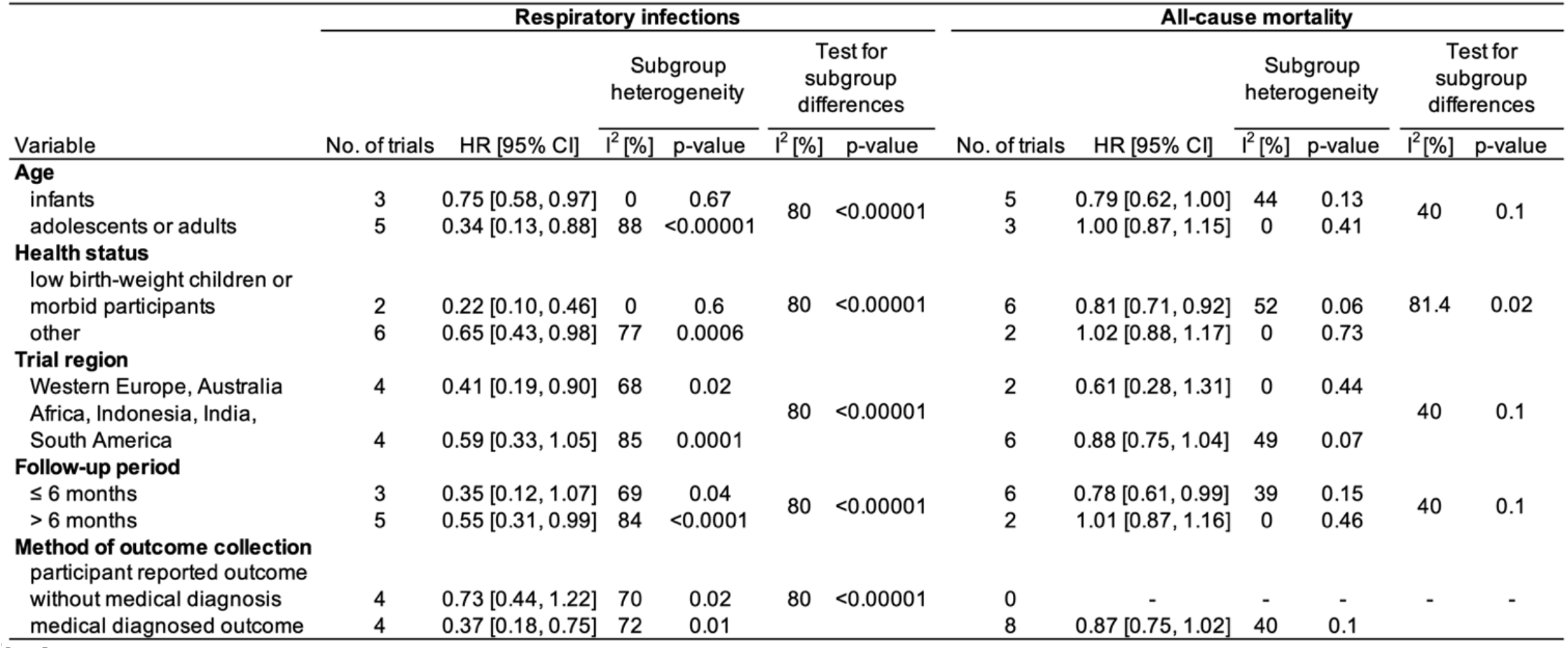
Subgroup analysis of BCG-trials for the outcomes respiratory infections and all-cause mortality. Outcomes were stratified by age, health status, trial region, follow-up period, and method of outcome collection. For trials conducted in infants and for COVID-19 related trials a follow-up period before the application of additional vaccines was used. HR = hazard ratio, CI = confidence interval

## Discussion

Our meta-analysis indicates that vaccination with BCG caused an estimated 49% decrease in risk for non-tuberculosis respiratory infections (HR 0.51, 95% CI 0.33-0.80). Further analysis revealed evidence for a protective effect after BCG vaccination of 22% on all-cause mortality if follow-up was restricted to one year (HR 0.78, 95% CI 0.63-0.96); there was no evidence for an effect when longer follow-up was considered (HR 0.87, 95% CI 0.75-1.02) [25]. In particular, mortality for infections (HR 0.67, 95% CI 0.46-0.99) and mortality for sepsis (HR 0.62, 95% CI 0.41-0.93) were significantly reduced after BCG-vaccination. For COVID-19 (HR 0.83, 95% CI 0.61-1.14), infections of any origin (HR 0.84, 95% CI 0.71-1.00), and sepsis (HR 0.82, 95% CI 0.62-1.07), evidence for protection after BCG vaccination was not statistically significant. Regarding hospitalization (HR 1.02, 95% CI 0.92-1.14), hospitalization for infections (HR 0.96, 95% CI 0.85-1.10), and hospitalization for respiratory infections (HR 0.82, 95% CI 0.31-2.18), we found no statistically significant improvement after BCG vaccination. Subgroup analyses suggest better protection of non-tuberculosis respiratory infections after BCG vaccination in adolescents or adults, in participants with impaired health including low-birth-weight children, in trials from Western Europe or Australia, in trials with a follow-up period shorter or equal to six months, and in trials with outcome collection by a medical diagnosis instead of participant-reported data. Regarding all-cause mortality, subgroup analyses indicate better protection in low-birth-weight children or morbid participants.

Based on the few RCTs included and the large number of potential effect modifiers and confounders, results of our subgroup-analysis must be interpreted with caution. The subgroup indicating less pronounced NSE for respiratory infections in infants includes only three RCTs [8,23,32]; two of them [8,33] collected outcome data by interviewing adults of participating infants. Diagnosing a respiratory infection in infants under 13 months of age might be challenging for an ordinary person without medical education, in particular as these two RCTs were originally designed to study the effect of BCG on allergies and therefore included a study population with a high degree of families with a history of atopy and asthma [34].

Therefore, allergic predisposition among infants might have influenced this outcome. Moreover, BCG vaccination causes a visible scar hindering blinding of participants and thereby introducing a possible source of bias. Results of our stratification by method of outcome collection support this speculation and suggest more pronounced BCG-mediated protective effects in trials with data collection based on a medical diagnosis. However, subgroup analysis of mortality, an objective outcome that is not affected by the method of outcome collection, indicates more pronounced protective effects in infants compared to adults.

The effect of routine childhood vaccinations on overall mortality was first systematically analyzed in observational studies in Guinea-Bissau. A large cohort study revealed that BCG vaccination was associated with a significantly lower mortality ratio of 0.55 (95% CI 0.36-0.85) [35]. These promising results lead to a systematic review commissioned by the WHO which identified five trials with an average relative risk of 0.70 (95% CI 0.49-1.01) and nine observational studies with an average relative risk of 0.47 (95% CI 0.32-0.69) by 2013.

Mortality reduction was most significant in two trials that were restricted to infants with low-birth-weight (RR 0.52, 95% CI 0.33–0.82) [3]. Another meta-analysis of three RCTs conducted in Guinea-Bissau showed that early BCG administration reduced mortality by 38% within the neonatal period (MRR 0.62, 95% CI 0.46–0.83) [6]. In 2014, the strategic advisory group of experts (SAGE) demanded a further confirmation of the results about NSE via the conduction of high quality RCTs [36].

Our meta-analysis is based on data from 15 RCTs reporting NSE of BCG; twelve of them were published after 2017 and to our knowledge not included in previous meta-analyses, five of them report mortality data. Based on this evidence, we did not find a significant reduction of all-cause mortality after BCG vaccination (HR 0.87, 95% CI 0.75-1.02). However, restriction to a follow-up of 12 months excluded one RCT [25], which contributed 43% weight to the overall analysis, and resulted in an overall significant protective effect in a range of previous findings (HR 0.78, 95% CI 0.63-0.96). This large RCT in Malawi [25] followed two populations for four and 16 years but could not detect any protective NSE after BCG vaccination. The authors concluded that the large number of non-infectious related deaths and the long time interval since BCG vaccination might have obscured any protective NSE.

The persistence of NSE of BCG has been analyzed in mechanistical studies demonstrating epigenetic reprogramming of monocytes, leading to increased cytokine production in response to non-related pathogens for up to three months after BCG vaccination, and enduring changes in pattern recognition receptors after 1 year. In healthy humans, non-specific Th1 and Th17 responses were enhanced for at least 1 year after vaccination [10]. In addition to this trained immunity, emergency granulopoiesis has been recently described as a mechanism providing non-specific protection within days of BCG-administration [12]. Moreover, heterologous T cell reactivity may also play a role [13]. Nevertheless, it is still unclear how long NSE after BCG vaccination last and whether they can be reversed.

Whereas previous meta-analysis quantified the impact of NSE of BCG on mortality, we focused our meta-analysis on non-tuberculosis respiratory infections and COVID-19.

Approximately 40 ongoing RCTs on this subject are a unique chance to elucidate the question whether and under which circumstances BCG protects from COVID-19. However, the launch of national COVID-19 vaccination programs interfered with ongoing BCG-COVID-19 trials. COVID-19 vaccinated people do not qualify for studies examining NSE of BCG alone on COVID-19. By the end of our search only four RCTs reporting data from participants not vaccinated with a COVID-19-specific vaccine were published and could be included in our analysis [15,26,27,31]. The number of people who are not willing to get vaccinated with a COVID-19 vaccine but volunteer for a BCG-COVID-19 trial is probably limited which impedes recruitment of study participants. Due to this fact it is questionable how many of these BCG-COVID-19 trials will be finished and when this data will be available. Our meta-analysis of published BCG-COVID-19 trials does not provide evidence for a protective effect of BCG-vaccination on COVID-19. Several effective COVID-19 specific vaccines exist by now. Nevertheless, the question of whether BCG can protect from respiratory infections in general is still highly relevant: 1) BCG’s ability to train the immune system and thereby protect antigen-independent against infections might provide an effective mean against vaccine-resistant mutants, which are able to circumvent antigen-specific immunological memory. Thus, BCG might be valuable to bridge time until an antigen-specific vaccine against a dangerous vaccine-resistant mutant is developed [37]. 2) BCG is safe, relatively inexpensive and can easily be provided on a mass scale [38]. According to current WHO policy, BCG is used for neonates in vaccination programs in developing countries in which the access to specific vaccines and medical supply is usually impaired [2]. This wide availability of BCG might be of strategic importance in pandemic preparedness: it could be easily provided and quickly be used for BCG-revaccination programs -with the potential to short-term prevent viral spread in these countries and therefore become a game-changer in the course of a future pandemic. 3) Antimicrobial resistance is a global health and development threat. It was declared by the WHO as one of the top 10 global public health threats facing humanity [40]. Globally, drug-resistant infections caused due to antimicrobial resistance contribute to about 700 000 deaths annually. Without effective intervention, drug-resistant infections are projected to cause 10 million deaths and a global economic loss of USD 100 trillion by 2050 (41). BCG-mediated training of the immune system might not only reduce the burden of infectious diseases but also decrease the need for antimicrobial prescriptions, thereby lowering the risk for the development of further drug-resistant pathogens.

## Conclusions

With rising resistance to antimicrobial interventions worldwide, improving the host response to infections becomes increasingly important. This meta-analysis summarizes the recent available RCT-data on BCG-mediated NSE and indicates time-limited and partial protection from respiratory infections. There was no evidence for a protective NSE of BCG-vaccination on COVID-19. Yet, the BCG-mediated NSE described here have the potential to short-term prevent viral spread and be of strategic importance in pandemic preparedness. In particular in developing countries with established BCG vaccination programs, BCG is easily accessible for revaccinations in pandemic emergencies which could positively influence the course of a future pandemic. Moreover, BCG-mediated NSE may reduce antimicrobial prescriptions and thus the development of antimicrobial resistance.

Our findings indicating a protective NSE on mortality within one year after BCG vaccination support the current WHO policy of providing BCG vaccination to all infants on the first day of life in areas of high infectious disease incidence [2]. However, as the setup of the included RCTs is heterogenic, the number of potential confounders and effect modifiers is large. There is a need for additional RCTs to clarify the circumstances under which BCG mediates NSE.

## Data Availability

All data produced in the present study are available upon reasonable request to the authors

## Acknowledgements

The authors would like to thank Beatrice Minder and Doris Kopp, information specialists at the Institute of Social and Preventive Medicine (ISPM), University of Bern, Switzerland, for excellent support for the development of the literature search strategy, and Prof. Taulant Muka, ISPM, University of Bern, Switzerland, for helpful feedback on intermediate data.

## Author contribution

G.T. was the lead investigator and responsible for setup of the research question and study.

G.T. and J.B. developed the final scientific protocol. G.T. and M.D. collected and evaluated the data. G.T. performed the statistical analysis. G.T. drafted the manuscript and coordinated manuscript revision. All authors provided critical evaluation and revision of the manuscript.

G.T. had final responsibility for the decision to submit for publication.

## Supporting Information

**S1 Fig. Funnel plots of random-effects meta-analysis of BCG trials for outcomes with at least eight studies**. A) Respiratory infections B) All-cause mortality C) All-cause hospitalization.

**S2 Fig. Forest plots of fixed-effect meta-analysis of BCG trials for infections of any origin**.

**S3 Fig. Forest plots of random-effects meta-analysis of BCG trials for A) hospitalization**

**B) hospitalization for infections C) hospitalization for respiratory infections**.

**S1 Table. Electronic data sources for systematic literature search**.

**S2 Table. Detailed search strategy used in this review**.

**S3 Table. Review checklist with in- and exclusion criteria**.

**S4 Table. Risk of bias in RCTs**.

**S5 Table. Reports excluded after assessment of full-texts for eligibility**.

