## Supplementary figures and images for "Non-specific effects of Bacillus Calmette-Guérin - a systematic review and meta-analysis of randomized controlled trials"

### S1 Fig

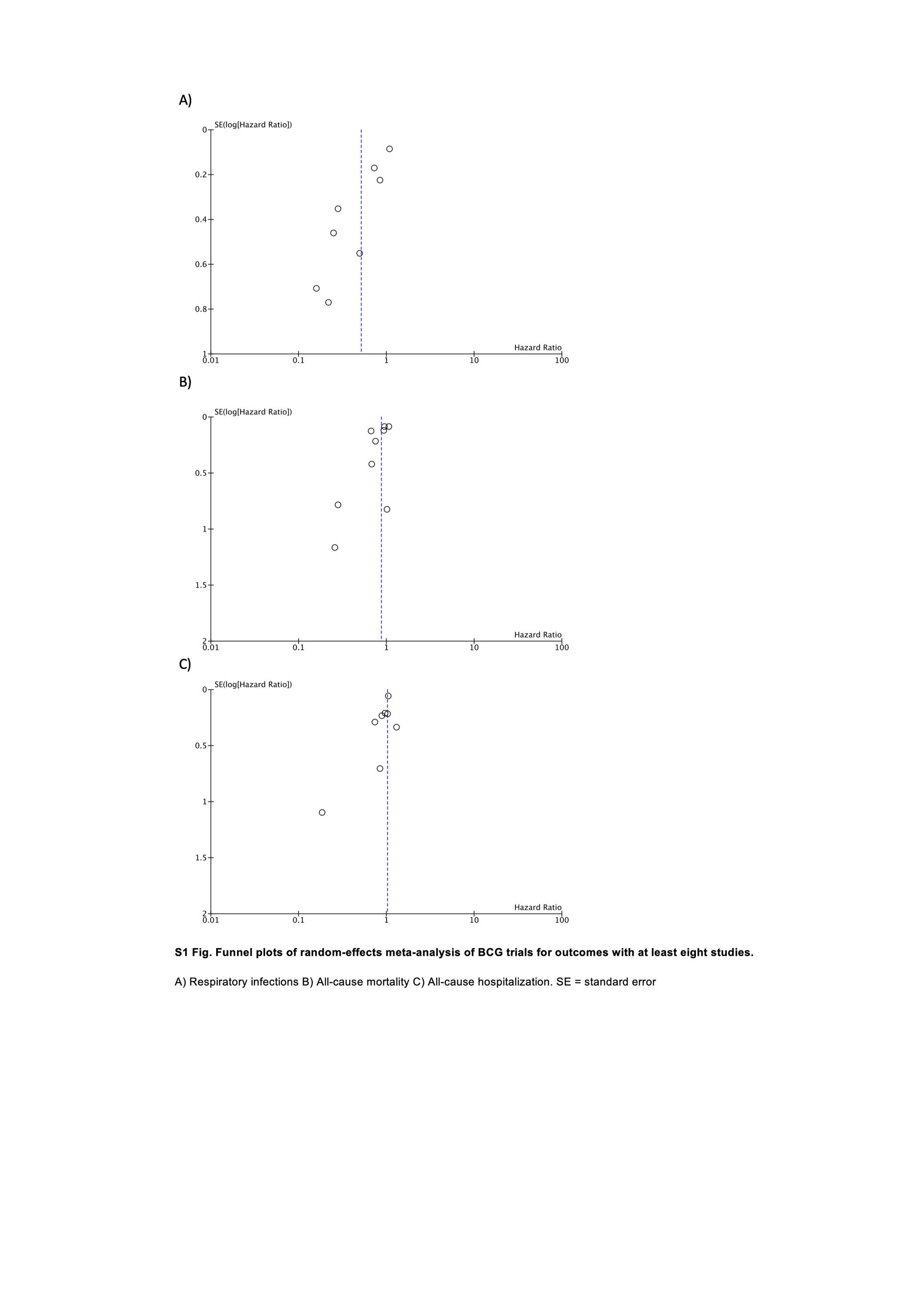

### S1 Table

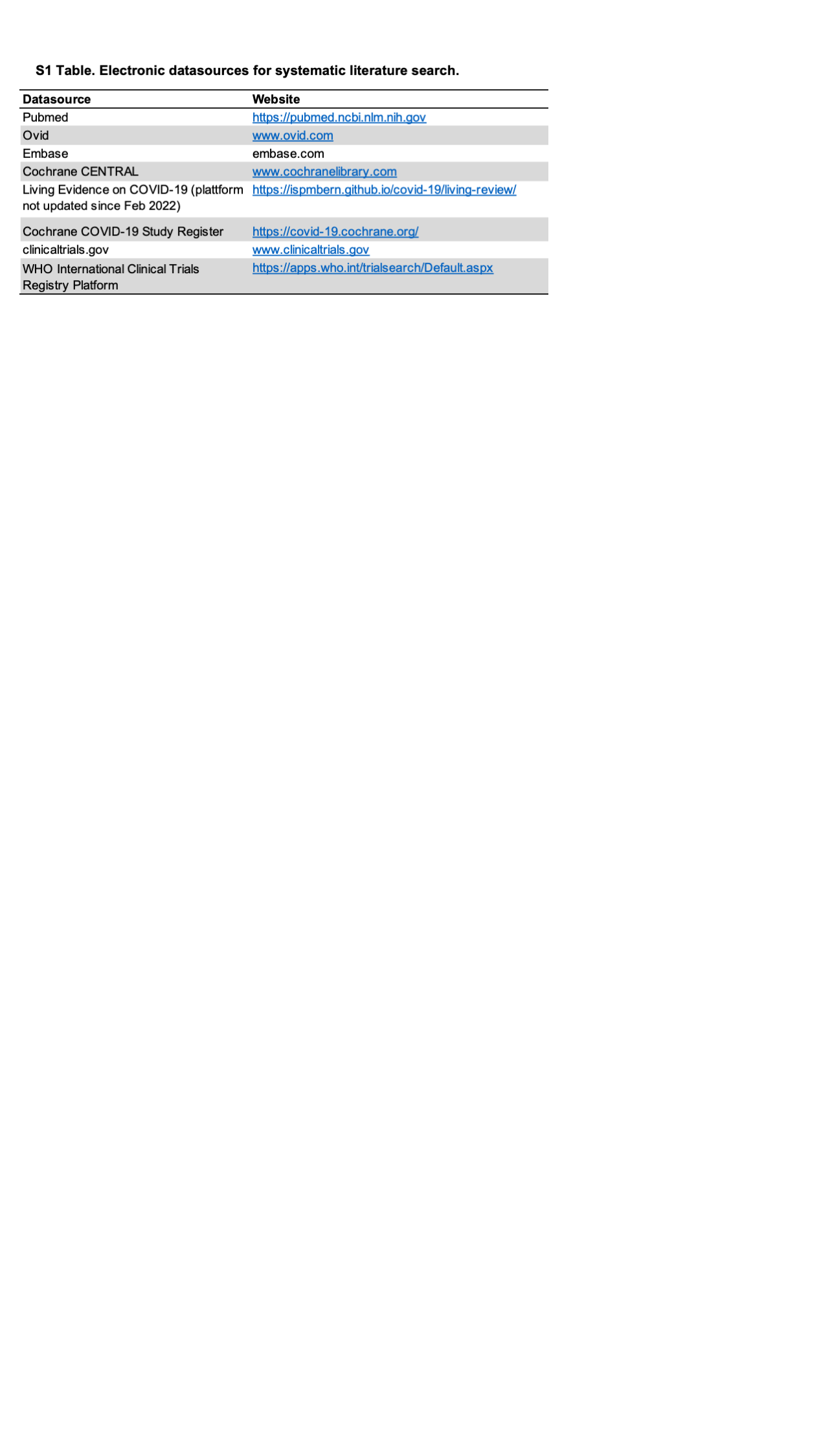

### S2 Fig

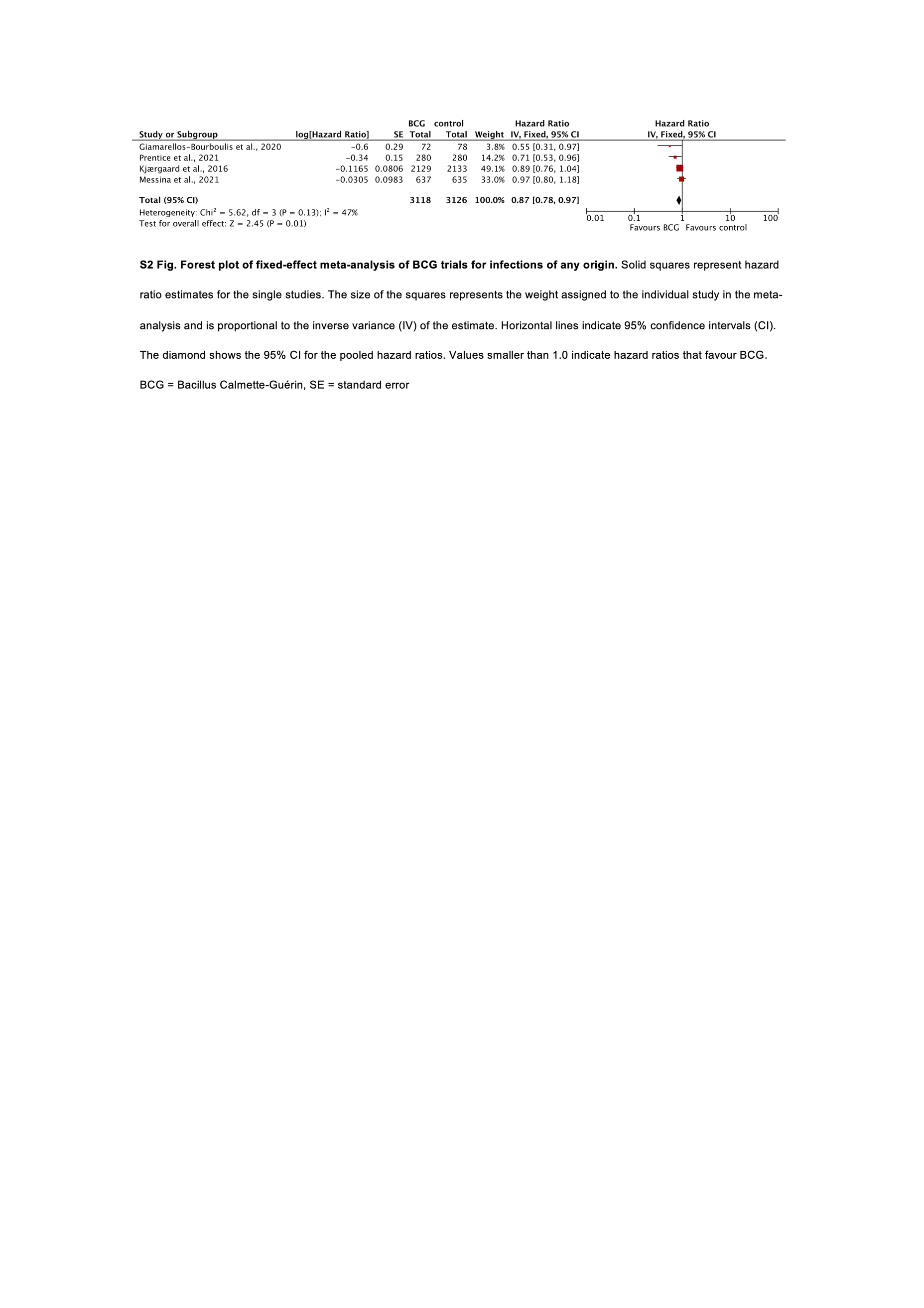

### S3 Fig

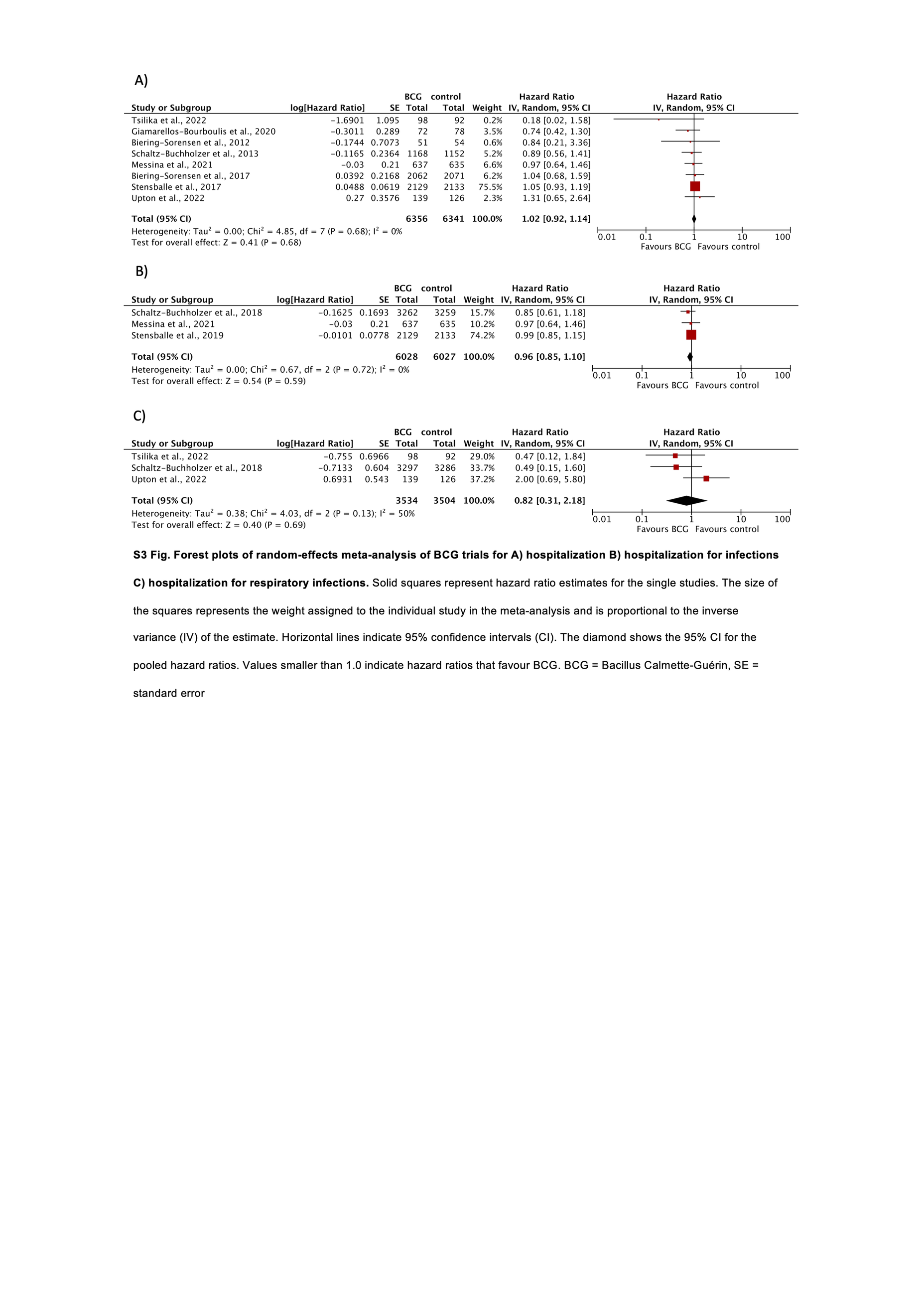

### S3 Table

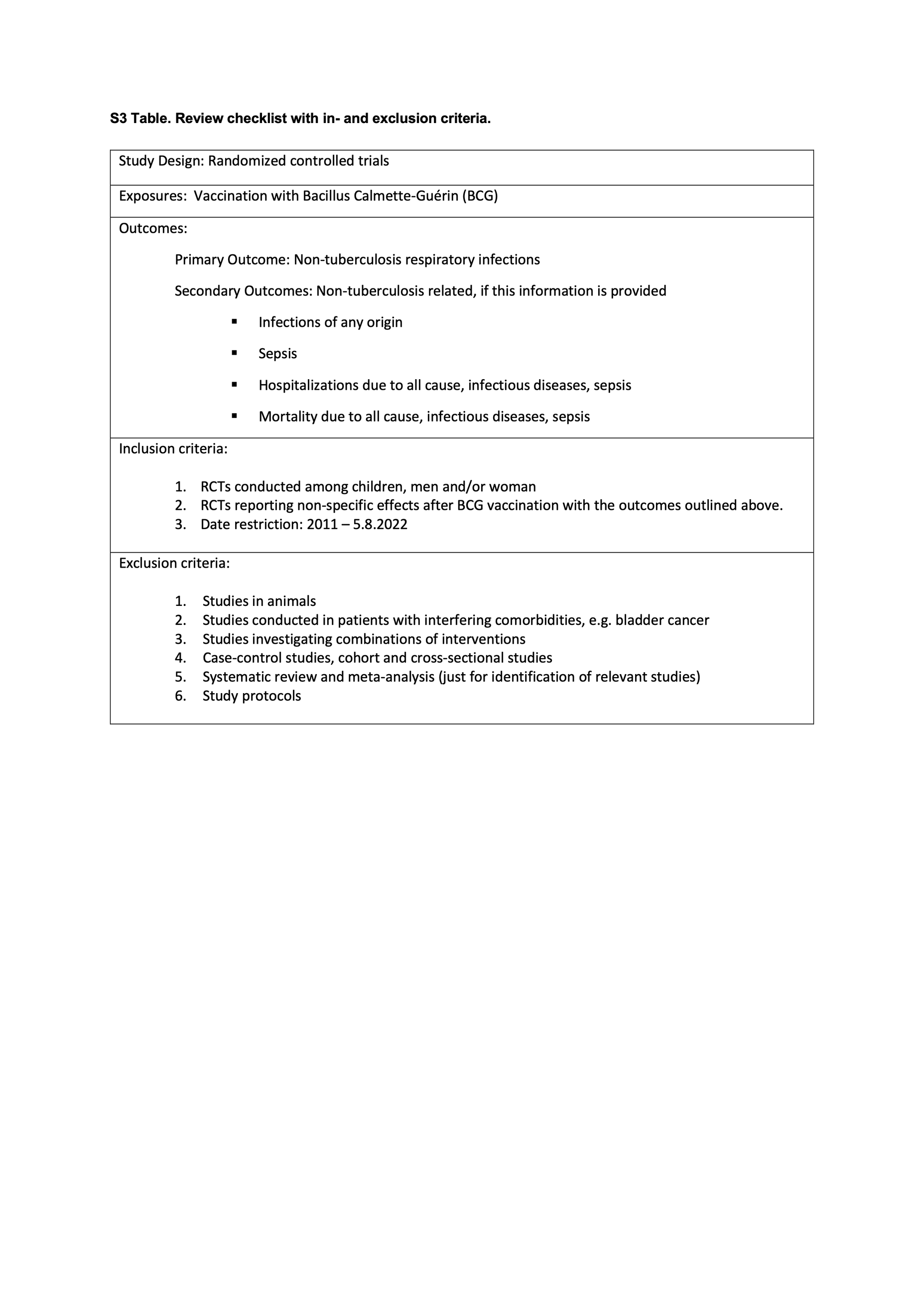

### S4 Table

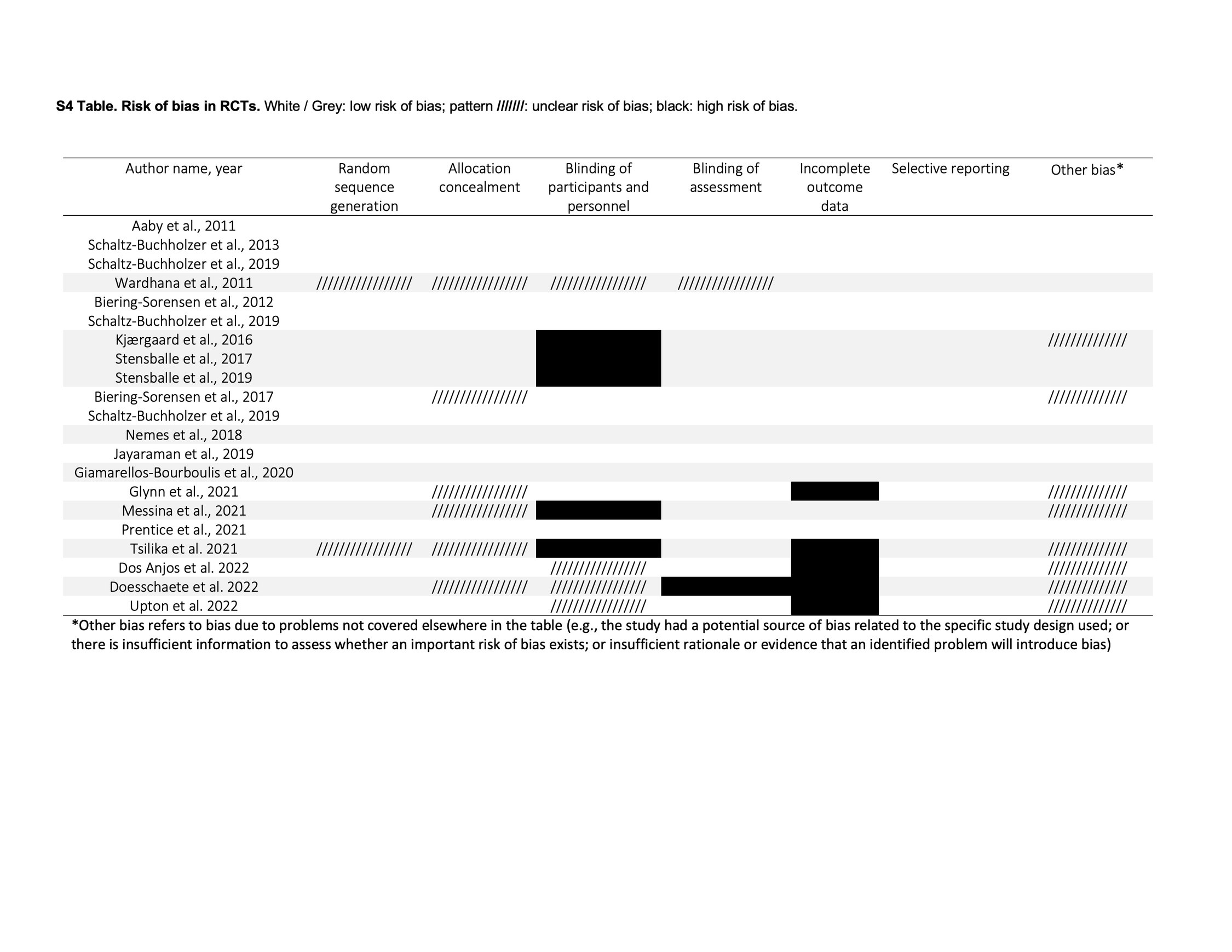
