## Supplementary material for "Non-specific effects of Bacillus Calmette-Guérin - a systematic review and meta-analysis of randomized controlled trials": S2 Table

**S2 Table. Detailed search strategy used in this review.**

| | |
|---|---|
| Pubmed | (tuberculosis vaccines[mh] OR bcg vaccine[mh] OR (aeras 402[tiab] OR ((antituberculosis[tiab] OR tuberculosis[tiab] OR tubercle bacilli[tiab]) AND (vaccin*[tiab] OR vacin*[tiab] OR vaksin*[tiab])) OR ((BCG[tiab] OR Calmette Guerin[tiab] OR Calmette*[tiab]) AND (vaccin*[tiab] OR vacin*[tiab] OR vaksin*[tiab] OR live[tiab] OR test[tiab] OR vacin*[tiab] OR bacillus[tiab] OR mycobacterium[tiab] OR copenhagen 1331[tiab])) OR immun bcg pasteur[tiab] OR immuno bcg pasteur[tiab] OR mva 85a[tiab] OR mva85a[tiab] OR mycobax[tiab] OR pastimmun[tiab] OR vpm[tiab] 1002[tiab] OR vpm1002[tiab])) AND (Respiratory Tract Infections[mh] OR COVID-19[mh] OR (((respiratory[tiab] OR respiration[tiab] OR airway[tiab] OR bronchopulmonary[tiab] OR pulmonary[tiab] OR pulmonal[tiab] OR lung[tiab] OR lungs[tiab] OR coronavir*[tiab]) AND (infect*[tiab] OR inflammat*[tiab] OR disease*[tiab] OR disorder*[tiab] OR illness*[tiab] OR syndrom*[tiab])) OR pneumoni*[tiab] OR covid 19[tiab] OR sars cov 2[tiab] OR 2019 ncov[tiab] OR ((new[tiab] OR novel[tiab] OR pandemic[tiab] OR epidemic[tiab]) AND (coronavirus*[tiab] OR corona virus*[tiab])) OR ((non-specific[tiab] OR nonspecific[tiab] OR heterologous[tiab]) AND (effect[tiab] OR effects[tiab])))) AND (randomized controlled trial[pt] OR controlled clinical trial[pt] OR random*[tiab] OR placebo[tiab] OR drug therapy[sh] OR randomly[tiab] OR trial[tiab] OR groups[tiab] NOT (animals [mh] NOT humans [mh]))<br><br>Filters applied: From 2011 to 2022 |
| Medline (Ovid) | (tuberculosis vaccines/ or bcg vaccine/ or (aeras 402 OR ((antituberculosis OR tuberculosis OR tubercle bacilli) ADJ3 (vaccin* OR vacin* OR vaksin*)) OR ((BCG OR Calmette Guerin OR Calmette*) ADJ3 (vaccin* OR vacin* OR vaksin* OR live OR test OR vacin* OR bacillus OR mycobacterium OR copenhagen 1331)) OR immun bcg pasteur OR mva 85a OR mva85a OR mycobax OR pastimmun OR vpm 1002 OR vpm1002).ab,ti,kf) AND (exp Respiratory Tract Infections/ OR COVID-19/ or (((respiratory OR respiration OR airway OR bronchopulmonary OR pulmonary OR pulmonal OR lung OR lungs OR coronavir*) ADJ3 (infect* OR inflammat* OR disease* OR disorder* OR illness* OR syndrom*)) OR pneumoni* OR covid 19 OR (sars ADJ2 cov 2) OR 2019 ncov OR ((new OR novel OR pandemic OR epidemic) AND (coronavirus* OR corona virus*)) OR ((non-specific OR nonspecific OR heterologous) ADJ3 (effect*))).ab,ti,kf) AND (exp Controlled clinical trial/ OR "Double-Blind Method"/ OR "Single-Blind Method"/ OR "Random Allocation"/ OR (random* OR factorial* OR crossover* OR (cross ADJ over*) OR placebo* OR ((doubl* OR singl*) ADJ blind*) OR assign* OR allocat* OR volunteer* OR trial OR groups OR rct).ab,ti.) AND not (exp animals/ NOT humans/) limit 12 to yr="2011 - 2022" |
| Embase | ('BCG vaccine'/de OR 'BCG vaccination'/de OR ('aeras 402' OR ((antituberculosis OR tuberculosis OR 'tubercle bacilli') NEAR/3 (vaccin* OR vacin* OR vaksin*)) OR ((b.c.g. OR BCG OR 'Calmette Guerin' OR Calmette*) NEAR/3 (vaccin* OR vacin* OR vaksin* OR live OR test OR vacin* OR bacillus OR mycobacterium OR 'copenhagen 1331')) OR 'immun bcg pasteur' OR 'mva 85a' OR mva85a OR mycobax OR pastimmun OR 'vpm 1002' OR vpm1002):ab,ti,kw) AND ('respiratory tract infection'/exp OR 'respiratory infections'/de OR 'Coronavirus infection'/exp OR 'covid 19'/exp OR 'coronavirus disease 2019'/exp OR (((respiratory OR respiration OR airway OR bronchopulmonary OR pulmonary OR pulmonal OR lung OR lungs OR coronavir*) NEAR/3 (infect* OR inflammat* OR disease* OR disorder* OR illness* OR syndrom*)) OR pneumoni* OR 'covid 19' OR (sars NEAR/2 'cov 2') OR '2019 ncov' OR ((new OR novel OR pandemic OR epidemic) AND (coronavirus* OR 'corona virus*')) OR ((non-specific OR nonspecific OR heterologous) NEAR/3 (effect*))):ab,ti,kw) AND ('clinical trial'/exp OR 'randomization'/de OR 'Crossover procedure'/de OR 'Double-blind procedure'/de OR 'Single-blind procedure'/de OR (random* OR factorial* OR crossover* OR (cross NEXT/1 over*) OR placebo* OR ((doubl* OR singl*) NEXT/1 blind*) OR assign* OR allocat* OR volunteer* OR trial OR groups OR rct):ab,ti) NOT ([animals]/lim NOT [humans]/lim) AND [2011-2022]/py |
| Cochrane CENTRAL | ("aeras 402" OR ((antituberculosis OR tuberculosis OR "tubercle bacilli") NEAR/3 (vaccin* OR vacin* OR vaksin*)) OR ((b.c.g. OR BCG OR "Calmette Guerin" OR Calmette*) NEAR/3 (vaccin* OR vacin* OR vaksin* OR live OR test OR vacin* OR bacillus OR mycobacterium OR "copenhagen 1331")) OR "immun bcg pasteur" OR "mva 85a" OR mva85a OR mycobax OR pastimmun OR "vpm 1002" OR vpm1002):ab,ti,kw AND (((respiratory OR respiration OR airway OR bronchopulmonary OR pulmonary OR pulmonal OR lung OR lungs OR coronavir*) NEAR/3 (infect* OR inflammat* OR disease* OR disorder* OR illness* OR syndrom*)) OR pneumoni* OR "covid 19" OR (sars NEAR/2 "cov 2") OR "2019 ncov" OR ((new OR novel OR pandemic OR epidemic) AND (coronavirus* OR corona NEXT virus*)) OR ((non-specific OR nonspecific OR heterologous) NEAR/3 (effect*))):ab,ti,kw<br><br>Year range: 2011-2022 |

| | |
|---|---|
| Cochrane COVID-19 Study Register | BCG OR Calmette* OR VPM1002<br><br>Limit: from date 01 /01 /2011 – 04 / 07 / 2022 |
| ClinTrials.gov | (BCG OR Calmette OR VPM1002 OR "vpm 1002" OR "copenhagen 1331" OR "immun bcg pasteur" OR "mva 85a" OR mva85a OR mycobax OR pastimmun) AND (respiratory OR respiration OR airway OR bronchopulmonary OR pulmonary OR pulmonal OR pneumonia OR lung OR lungs OR coronavirus OR "corona virus" OR covid OR SARS-Cov-2 OR ncov) |
| WHO International Clinical trials Registry Platform | (BCG OR Calmette OR VPM1002 OR "vpm 1002" OR "copenhagen 1331" OR "immun bcg pasteur" OR "mva 85a" OR mva85a OR mycobax OR pastimmun) |
| Living Evidence on COVID-19 database | (BCG) OR (Calmette) OR (VPM1002) OR (mva85a) OR (mycobax) OR (pastimmun) |
