## Supplementary material for "Non-specific effects of Bacillus Calmette-Guérin - a systematic review and meta-analysis of randomized controlled trials": S5 Table

**S5 Table. Reports excluded after assessment of full-texts for eligibility.**

| Reason for exclusion | Lead author, publication date | Link to publication |
|---|---|---|
| Relevant outcomes not reported | Messina et al., 2022 | https://doi.org/10.1002/cti2.1387 |
| | Pittet et al., 2022 | DOI: 10.1016/j.vaccine.2022.01.005 |
| | Thostesen et al., 2016 | http://dx.doi.org/10.1111/all.12969 |
| | Saini et al., 2021 | DOI: 10.1183/13993003.congress-2021.PA450 |
| Study design not relevant | Quinn et al., 2022 | https://doi.org/10.1016/j.vaccine.2022.04.082 |
| | Amirlak et al., 2020 | https://doi.org/10.1101/2020.08.10.20172288; |
| | Leentjens et al., 2015 | DOI: 10.1093/infdis/jiv332 |
| | Tameris et al., 2019 | http://dx.doi.org/10.1016/S2213-2600(19)30251-6 |
| Background article | Gong et al., 2021 | DOI: 10.1080/14760584.2021.1938550 |
| Application of additional vaccines | Moorlag et al., 2022 | doi: 10.1093/cid/ciac182 |
| | Czajka et al., 2022 | https://doi.org/10.3390/vaccines10020314 |
| | Schaltz-Buchholzer et al., 2021 | DOI: 10.1093/infdis/jiab220 |
| | Villanueva et al., 2022 | https://doi.org/10.1371/journal.pone.0268042 |
| | Ramos-Martinez et al., 2021 | doi: 10.3390/cells10113179 |
